# Temporal Analysis of Social Determinants Associated with COVID-19 Mortality

**DOI:** 10.1101/2021.06.22.21258971

**Authors:** Shayom Debopadhaya, John S. Erickson, Kristin P. Bennett

## Abstract

This study examines how social determinants associated with COVID-19 mortality change over time. Using US county-level data from July 5 and December 28, 2020, the effect of 19 high-risk factors on COVID-19 mortality rate was quantified at each time point with negative binomial mixed models. Then, these high-risk factors were used as controls in two association studies between 40 social determinants and COVID-19 mortality rates using data from the same time points. The results indicate that counties with certain ethnic minorities and age groups, immigrants, prevalence of diseases like pediatric asthma and diabetes and cardiovascular disease, socioeconomic inequalities, and higher social association are associated with increased COVID-19 mortality rates. Meanwhile, more mental health providers, access to exercise, higher income, chronic lung disease in adults, suicide, and excessive drinking are associated with decreased mortality. Our temporal analysis also reveals a possible decreasing impact of socioeconomic disadvantage and air quality, and an increasing effect of factors like age, which suggests that public health policies may have been effective in protecting disadvantaged populations over time or that analysis utilizing earlier data may have exaggerated certain effects. Overall, we continue to recognize that social inequality still places disadvantaged groups at risk, and we identify possible relationships between lung disease, mental health, and COVID-19 that need to be explored on a clinical level.

**CCS CONCEPTS:**

- **Applied computing** → **Health informatics**.

## 1 INTRODUCTION

The growing prevalence of COVID-19 has forced the pandemic to be the center of state and national policy in the United States (US). As of July 5, 2020, the CDC documented over 130,000 COVID-19 deaths in the US [24], and this number increased to over 300,000 COVID-19 deaths by December 28, 2020 [62]. This increase in deaths is not uniformly distributed, and several counties are experiencing much higher-than-average death rates [49]. It is hypothesized that social determinants have contributed to these disparities in COVID-19 mortality [1], which we investigate. Social determinants, such as a county’s access to healthcare, rates of education, indicators of health, and economic status have greatly impacted other diseases [14], so these factors may play a similar role in COVID-19.

An increasing amount of literature highlights that social determinants of COVID-19 place socioeconomically disadvantaged populations at heightened risk. Abrams and Szefler reviews emerging literature to discuss the stark inequality of COVID-19 infection rates and outcomes among several groups [1]. This work recognizes that social determinants such as housing problems, race, smoking, nutrition, overcrowding, poverty, and comorbidities may compromise an individual to COVID-19. Ahmed et al. is another among the many to describe the disproportionate effects of COVID-19 among the socioeconomically disadvantaged, highlighting access to health-care as one of the many determinants that are potentially critical in COVID-19 mortality [2]. Fielding-Miller et al. studies a few select social determinants to find that dense population in urban counties, non-English speakers, farm workers, and impoverished groups in non-urban counties are at increased risk [22]. Overall, the literature emphasizes that quantifying the social determinants of COVID-19 is a crucial step in addressing the existing health inequalities. Yet, there is a lack of well-controlled, diverse screening of the social determinants associated with COVID-19. Moreover, as the COVID-19 pandemic is rapidly changing, temporal analysis is needed to understand how the effects of these socioeconomic factors respond to the implementation of systemic protections.

To address the gap in the current literature and leverage readily available data, this study identifies and compares which social determinants are associated with county-level changes in COVID-19 mortality rates on July 5 and December 28, 2020. Through this temporal analysis, we uniquely utilize a comprehensive list of co-morbidities, social determinants, and the impact of differing state policies to explore the nationwide effect of socioeconomic disparities.

## 2 METHODS

This study consists of an initial ecological analysis to establish which high-risk factors from literature have statistically significant relationships to COVID-19, using pandemic mortality data updated to July 5 and December 28, 2020. Then, the high-risk factors are used as controls for two social determinant association studies - one using the COVID-19 mortality data updated until July 5 and one using COVID-19 mortality data updated until December 28. These dates were selected to avoid the volatile increase in COVID-19 infections in the two weeks following major US holidays [41]. 40 social determinants are evaluated at each time point to find which ones affect COVID-19 mortality. Both the two initial analyses and the two follow-up association studies utilize negative binomial mixed models to analyze county level data (n=3093 counties). Results are statistically corrected for possible false discoveries. Further sensitivity analysis with different model variants and additional time-series analysis are also validated. This study design is summarized in Figure 1.

**Figure 1:**
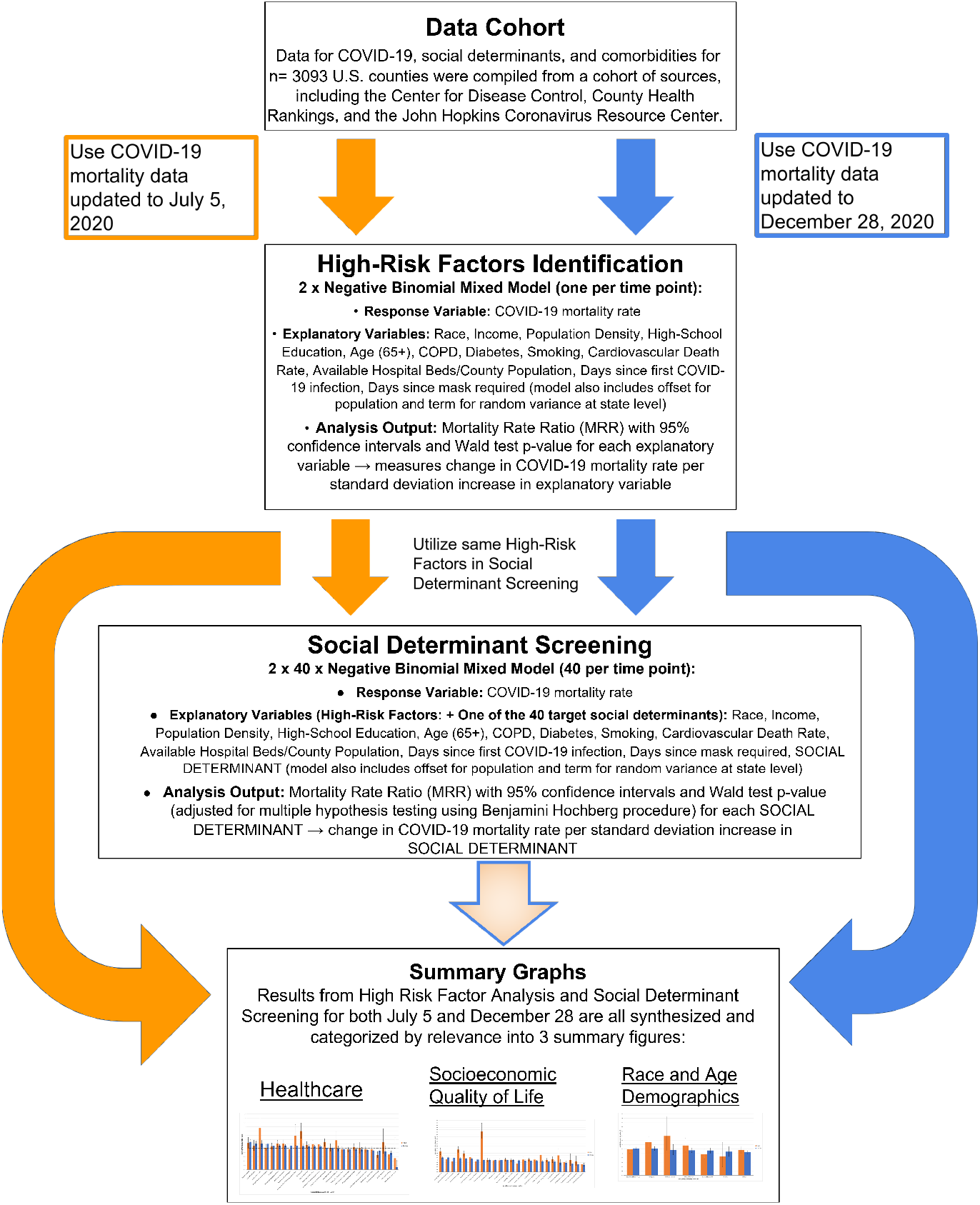
Study Design Flowchart. The chart shows the methodology from the data aggregation in the Data Cohort, to the initial ecological analysis in the High-Risk Factors Identification stage, use of High-Risk Factors in the Social Determinant Screening, and combination of High-Risk Factors analysis and Social Determinant Screening results into Summary Graphs. The orange arrows on the left denote the use of COVID-19 mortality data updated until July 5, 2020. The blue arrows on the right denote the use of COVID-19 mortality data updated until December 28, 2020. The larger orange and blue curved arrows and the mixed blue border/orange gradient arrow indicates how high-risk factors and social determinants for both dates are synthesized in the Summary Graphs.

County-level COVID-19 mortality data on July 5 and December 28, 2020 was sourced from John Hopkins University Center for Systems Science and Engineering [19]. While COVID-19 mortality rates change over the course of this study, the data for high-risk factors and social determinants are long-term characteristics of a county and remain constant throughout this study. Data for high-risk factors and social determinants were sourced from the US Census[10], County Health Rankings [54], JHU-CSSE [19], the Center of Disease Control (CDC) Wonder [67], American Lung Association [6], and several other data sources. The same risk factor and social determinants datasets were used for the July 5 and December 28, 2020 analyses since they were measured before the pandemic. All data is publicly accessible, and all methods and results are reproducible. The full R code and a supplementary document with complete list of data sources can be found on the Rensselaer IDEA Github^1^.

### 2.1 Identifying High-Risk Factors For COVID-19 using July 5 and December 28, 2020 Data

A robust set of controls needed to be established before screening for social determinants [40]. To find the most important risk factors, we considered the guidelines from the CDC. Using extensive meta-analysis, the CDC recognized that those suffering from several diseases and ethnic minorities are at a higher risk for severe illness and death due to COVID-19 [26]. Our analysis accounts for these well-documented risk factors, using available data for the racial distribution in the US [10], as well as data for several of the highest risk comorbidities, such as the prevalence of cardiovascular death, diabetes, and COPD [6, 54, 67]. Income and education levels were used to briefly explore socioeconomic status [2]. A breakdown of population density by quartile was used as a proxy to categorize the urban or rural identity of a county. Additionally, temporal variables such as days since first infection and days since a mask is required were included to adjust for disease progression and emerging policy in each state, as this would greatly affect mortality rates of any region [19]. As several other metrics of policy were previously tested and had little significance in aggregate data, they were omitted from this analysis [18]. Variables were scaled by subtracting the mean from each data point and dividing by the standard deviation.

Using county-level data for the entire U.S. (n=3093), negative binomial mixed models were used to find the significant high-risk factors. The modeling method was a generalization of those used in a prior study on the impact of air pollution on COVID-19 mortality [68]. The county COVID-19 mortality rate was used as the observed variable. The following explanatory variables were used: percent of county that is African American, percent of county that is Hispanic, percent of county that is Native American, percent of county that is Asian, percent of county that is White, percent of county with less than a high-school education, percent of county that is above the age of 65, percent of county that has diabetes, percent of county with COPD, percent of adults in the county who smoke, population density in quartiles, days since first infection, days since mask required, available hospital beds per county population, median household income of a county, and cardiovascular death rate. An offset to scale for population and a term to account for random variance at the state level was also included [18, 68, 69]. This analysis was performed twice, using COVID-19 mortality data updated up to July 5, 2020 and up to December 28, 2020.

The extracted output from this model is the the ratio of change in COVID-19 mortality rate per 1 standard deviation increase in each explanatory variable. We refer to this output as Mortality Rate Ratio (MRR) [68]. In this initial analysis of high-risk factors, these explanatory variables reveal the important baseline effects that must be controlled for when screening for social determinants.

### 2.2 Screening for Social Determinants

The same high risk factors from the methods above were used as controls for the two social determinant screenings. This study design allows for the identification of unique social determinants that are distinguishable from the already recognized high-risk factors.

For the social determinant screening, 40 negative binomial mixed models (each with high-risk factors as controls) were used in the association analysis to individually test 40 variables that represent a wide range of social determinants. This set of determinants included metrics of a county’s mental health, physical health, rates of disease, economic status, housing burdens, education, demographics, and multiple death rates, sourced from County Health Rankings [54]. The analysis produces a MRR for each of the 40 determinants at each of the 2 time points. A complete list of the tested explanatory variables and their sources can be found in the supplementary document.^1^

This analysis was performed using the COVID-19 mortality data from both July 5, 2020 and December 28, 2020, producing two sets of statistically significant social determinants associated with COVID-19 mortality.

### 2.3 Statistical Testing

Each explanatory variable in the high-risk models and each variable in the 40 social determinant association models were tested for statistical significance using a Wald test [66]. The Benjamini-Hochberg Procedure [7] was used to adjust p-values for Multiple Hypothesis Testing in the social determinant screening. Our models showed robustness in this process, as the false discovery rate was acceptably below 0.05 at 0.0297 and 0.012 for July 5 and December 28 models, respectively. A 95% confidence interval was also produced for each term’s MRR. The 95% confidence interval and the p-value is indicated displayed on each bar on Figures 2, 3, and 4.

**Figure 2:**
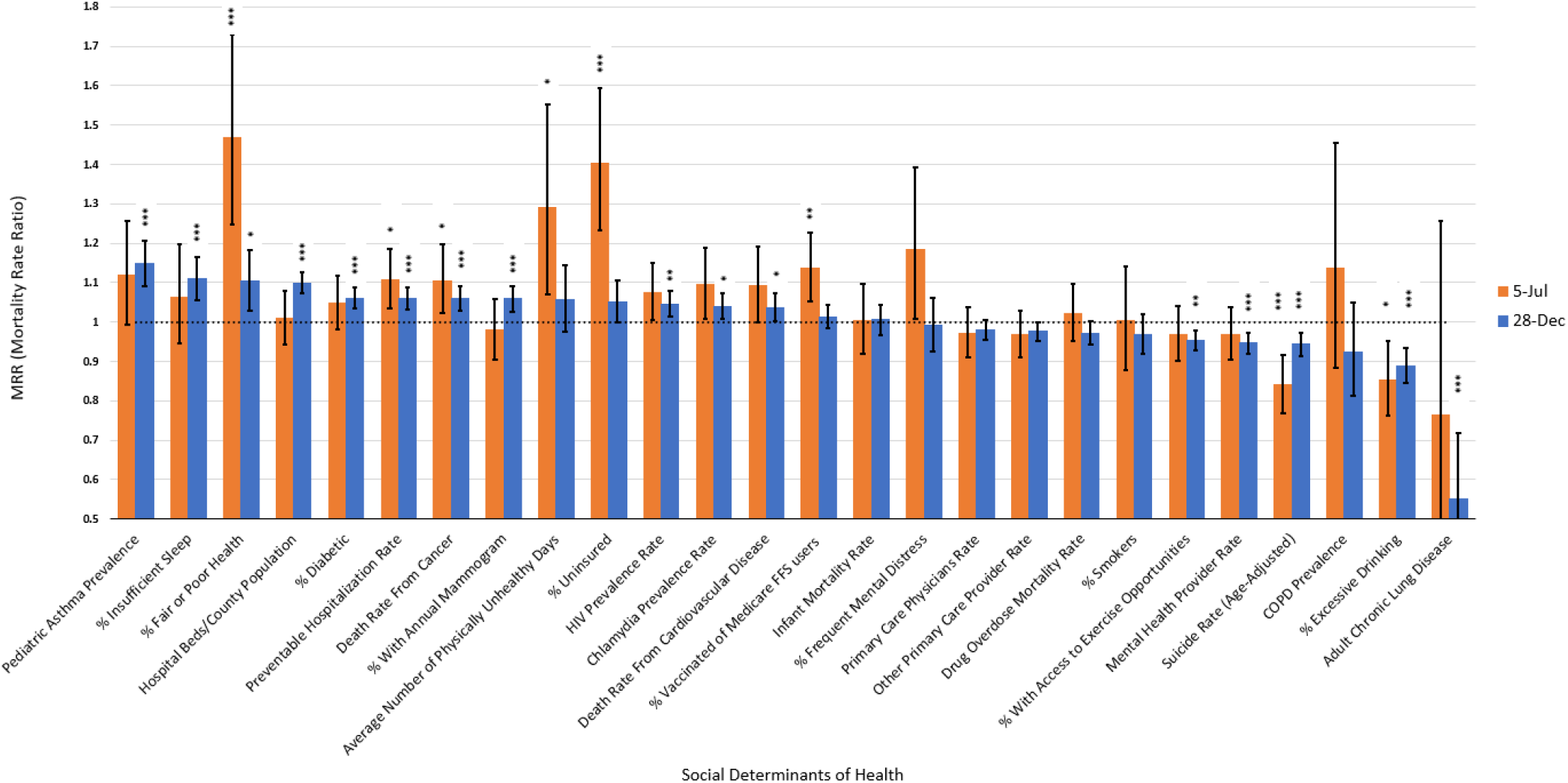
Mortality Rate Ratios Associated with Healthcare. The vertical axis shows Mortality Rate Ratio. The horizontal axis lists terms related to healthcare. The bars indicate a term’s respective MRRs at each time point. Bars are colored in orange to indicate analysis from July 5, 2020 data or colored in blue to indicate analysis from December 28, 2020 data. A dotted horizontal line indicates an MRR of 1, which represents no change in mortality rate. Each bar is labelled with its 95% confidence intervals and statistically significant, Benjamini-Hochberg corrected p-values are indicated as follows: * = p<0.05; ** = p<0.01; *** = p<0.001.

**Figure 3:**
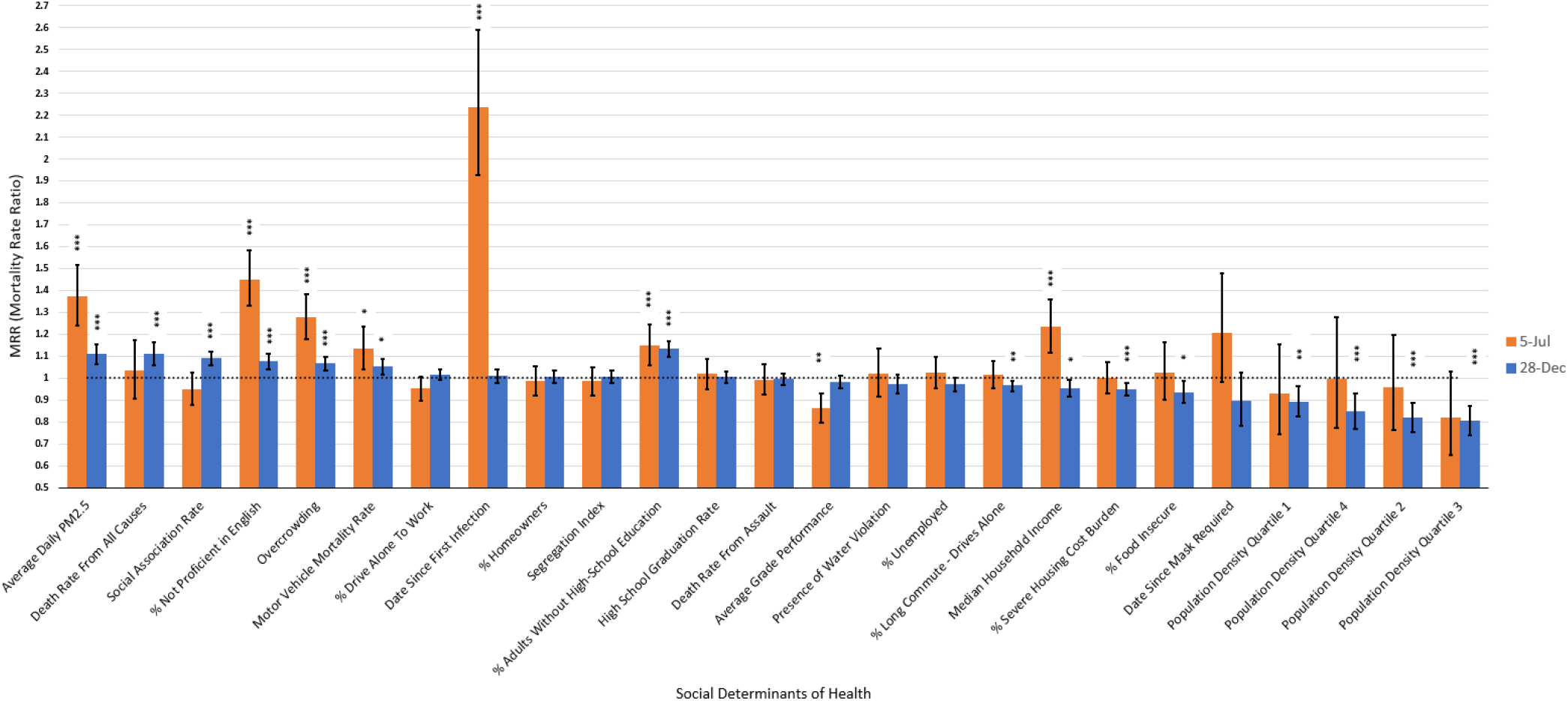
Mortality Rate Ratios Associated with Socioeconomic Quality of Life. The vertical axis shows Mortality Rate Ratio. The horizontal axis lists terms related to socioeconomic and regional characteristics. The bars indicate a term’s respective MRRs at each time point. Bars are colored in orange to indicate analysis from July 5, 2020 data or colored in blue to indicate analysis from December 28, 2020 data. A dotted horizontal line indicates an MRR of 1, which represents no change in mortality rate. Each bar is labelled with its 95% confidence intervals and statistically significant, Benjamini-Hochberg corrected p-values are indicated as follows: * = p<0.05; ** = p<0.01; *** = p<0.001.

**Figure 4:**
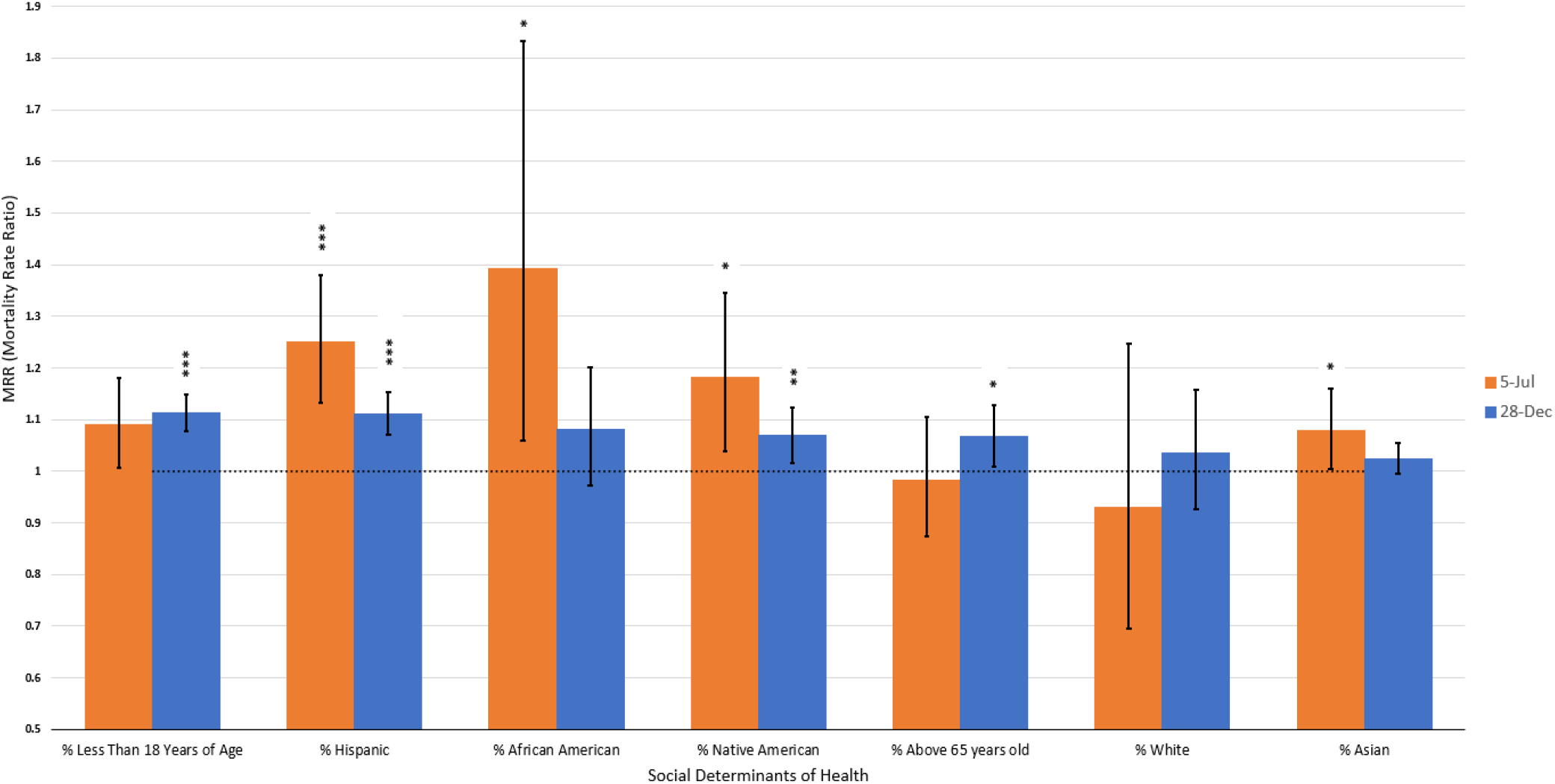
Mortality Rate Ratios Associated with Race and Age Demographics. The vertical axis shows Mortality Rate Ratio. The horizontal axis lists terms related to race and age demographics. The bars indicate a term’s respective MRRs at each time point. Bars are colored in orange to indicate analysis from July 5 or colored in blue to indicate analysis from December 28. A dotted horizontal line indicates an MRR of 1, which represents no change in mortality rate. Each bar is labelled with its 95% confidence intervals and statistically significant, Benjamini-Hochberg corrected p-values are indicated as follows: * = p<0.05; ** = p<0.01; *** = p<0.001.

To test performance and fit, the procedure from Wu et al. was followed [68]. The negative binomial mixed model for the high-risk factors using July 5, 2020 data was compared to zero inflated, fixed negative binomial, and spatial correlation variants. These models were compared to the negative binomial mixed model using AIC and BIC. The main high-risk model was also analyzed at 16 different time points for robustness. Additionally, variants of the model that excluded New York city data and excluded counties with less than 10 cases were analyzed for robustness. Statistical variations produced no significantly different results. Model was also robust to short-term time-series analysis. Any changes in results due to different sample stratification can be found in the supplementary document. All analysis was conducted in R [48] using the lme4 package, and visualizations were created using Microsoft Excel. Notebooks to reproduce all results can be found on the Rensselaer IDEA Github.

## 3 RESULTS

The results from the high risk analyses on July 5 and December 28 data were aggregated with the results from the two social determinant screenings. Then, this combination of analysis was categorized by relevance into 3 distinct sections for temporal comparison: “Healthcare”, “Socioeconomic Quality of Life”, and “Race and Age Demographics”. For each of the corresponding section figures, the MRR of each term is presented as a multiple of 1. A MRR above 1 indicates a predicted increase in mortality, and a value below 1 indicates a predicted decrease in mortality. A term is considered statistically significant if its 95% confidence interval does not cross the threshold of MRR of 1.

### 3.1 Healthcare

The Healthcare category combines the prevalence of smoking and prevalence of COPD from the high-risk analyses with several social determinants related to quality of healthcare and other metrics of population health. These results are presented in Figure 2.

### 3.2 Socioeconomic Quality of Life

The Socioeconomic Quality of Life category combines the median household income, population density, education (percent of adults in county without high-school education), days since first infection, and days since mask required variables from the high risk factors analyses with several social determinants that define a county’s living and working conditions. These results are presented in Figure 3.

### 3.3 Race and Age Demographics

The Race and Age Demographics category combines the racial composition of a county and percent of county above 65 years old from the high risk analyses with the social determinant of percent of county that is less than 18 years of age. These results are presented in Figure 4.

## 4 DISCUSSION

This study conducts temporal analysis of social determinants and comorbidities at the national scale with a wide variety of controls and county level data from July 5 and December 28, 2020. We reveal the high-risk factors and the specific social determinants of a county that affect the mortality rate of COVID-19.

### 4.1 Healthcare

This section discusses the results presented in Figure 2.

#### 4.1.1 Reduced Access to High Quality Healthcare

On July 5, one standard deviation increase in preventable hospitalization rate and percent of uninsured population was associated with a 1.11 and 1.40 times increase in COVID-19 mortality rate, respectively (1.11 and 1.40 MRR). By December 28, only preventable hospitalization rate had statistical significance (1.06 MRR). These social determinants are direct indicators of quality of healthcare. Overuse of hospitals typically indicates that outpatient care is inadequate or that there is limited available primary care [53]. Counties that were ill-equipped for their population pre-pandemic may continue to be inadequate to their populations during a pandemic, leading to more COVID-19 deaths. A high percent of uninsured individuals also indicates poor access to health care. In general, those without access to health insurance will not be given thorough preventative care, often resulting in undiagnosed health problems or severe illness [30], which may result in more COVID-19 deaths. As these determinants were found to have an MRR above 1, we recognize that poor healthcare is associated with increased COVID-19 mortality.

One standard deviation increase in average number of physically unhealthy days in the county and percent of county population with poor or fair health was associated with a 1.29 and 1.47 COVID-19 MRR on July 5, respectively. By December 28, only the effect of percent of county with poor or fair health remained statistically significant (MRR of 1.10). These findings suggest that counties with poor health outcomes before the pandemic have poor COVID-19 outcomes. Unhealthy counties may have a high prevalence of common underlying health conditions, which places them at increased risk to COVID-19, as recognized by the CDC [51]. Poor access to healthcare also has a likely role in the increased COVID-19 mortality rate in these counties, as poor mental and physical health has been linked to socioeconomic disadvantage [65]. However, it is possible that the decreasing statistical significance for physically unhealthy days and the decreasing MRR of poor or fair county health indicate that the effect of socioeconomic disadvantage and poor healthcare decreases throughout the pandemic.

One standard deviation increase in a county’s cancer death rate was associated with a 1.11 times increase in COVID-19 death rate on July 5, 2020. This MRR decreased to 1.06 by December 28. Higher cancer death rate can result both from increased incidence and unfavorable outcomes due to poor access to healthcare. High cancer mortality rates indicate that a community has insufficient prevention, early diagnosis, and treatment [60]. Low income communities are notably at a disadvantage for cancer treatment due to a lack of resources, as wealth disparities are noted as the most common cause of health disparities [4]. We recognize cancer prevalence as a likely comorbidity and predictor of poor healthcare, which increases risk of COVID-19.

#### 4.1.2 Suicide, Mental Health Provider Rate, and Excessive Drinking

We observed a statistically significant, negative relationship (MRR<1) between age-adjusted suicide rate and excessive drinking with COVID-19 mortality rate (0.84 and 0.85 MRR by July 5, respectively). The impact of these effects approached an MRR of 1 by December (0.94 and 0.89 MRR, respectively). Meanwhile, the relationship between mental health provider rate and COVID-19 mortality, which is statistically insignificant in July, has a statistically significant MRR of 0.95 in December. This suggests a complex relationship between the pandemic and the recent epidemic of “deaths of despair”: deaths from suicide, overdose, alcoholism, and self-harm. Alcohol is a coping strategy for mass stress and isolation, and consumption increased greatly after the pandemic began [38]. Suicide rates are higher in less urban areas and lower in more urban areas while the reverse is true for COVID-19 mortality rates [37]. While supporting effective social distancing, social isolation is a major risk factor for suicide [11]. The negative relationship of suicide and alcohol consumption with COVID-19 deaths are likely only temporary as suicide rates and alcohol abuse are likely to increase during the pandemic [34]. However, the emergence of higher mental health provider rates (indicating better access to mental health care) as a protective determinant suggests that mental health has a non-trivial, intricate relationship with COVID-19, and further work is needed to fully understand what we observe.

#### 4.1.3 Vaccination Rates and Annual Mammogram Rates in Medicare FFS

Influenza vaccination rates among Medicare Fee For Service (FFS) users was determined to have a 1.14 MRR in July but was not significant in December. Inversely, annual mammogram rates among Medicare FFS users was statistically insignificant in July but had a 1.06 MRR in December. One explanation for this result is that elderly groups in community care and those with pre-existing conditions like cardiovascular disease vaccinate and get screened for breast cancer more frequently. 83.1% of Medicare FFS users are above the age of 65 [43], and flu vaccinations are more common in individuals above 65 years old (72.3%) compared to those between 18 and 64 years old (30.7%) [23]. Flu vaccination rates are also higher in patients with comorbidities like cardiovascular disease in comparison to patients without cardiovascular disease [3]. Similarly, as there is an increased risk of breast cancer with age, increased mammogram screening among FFS users may represent elderly populations and groups with comorbidities [25]. These groups are more vulnerable COVID-19 [26], so the overall observed relationship between mammogram and vaccination rates in FFS users may reflect an underlying relationship between at-risk groups and COVID-19. However, as there are severe geographical, racial, and socioeconomic disparities in influenza vaccination rates [35], future analysis of vaccination, mammogram rates, and COVID-19 mortality is needed to reveal more about this complex relationship.

#### 4.1.4 Lung Disease

Surprisingly, one standard deviation increase in adult chronic lung disease in a county was associated with a statistically significant decrease in COVID-19 mortality rate in December (0.76 MRR) but was not significant in July. Rather than protective, the CDC identified several lung diseases (asthma, COPD, lung cancer) as a possible COVID-19 comorbidity [28]. While previous literature has observed this effect in asthma, this effect was not previously reported in other lung diseases [18]. As the tested COPD factor has no statistical significance at July or December, it suggests that lung disease other than COPD may have an increased statistical importance. Moreover, pediatric asthma was observed to have the opposite effect to adult chronic lung disease. Pediatric asthma was statistically insignificant on July 5, but was associated with a 1.15 MRR by December 28. As increased transmission of COVID-19 has been implicated in youth, the predicted increase in mortality from pediatric asthma may indicate a risk from greater youth populations in a county [20]. Overall, further analysis with specific lung disease subtypes is needed to better characterize the observed associations with lung disease.

#### 4.1.5 Sexually Transmitted Diseases (STDs)

One standard deviation increase in chlamydia prevalence rate and HIV prevalence rate was associated with a 1.09 and 1.07 times increase in COVID-19 mortality rates by July 5. By December, chlamydia and HIV prevalence had MRRs of 1.04 and 1.05. Chlamydia prevalence has been linked to minority women populations, and is related to several increases in mortality like cervical cancer and various inflammatory conditions [15, 32]. HIV prevalence is an indirect metric of high-risk behaviors such as unsafe sex and intravenous drug use [29]. Both HIV and chlamydia prevalence also indicate significant burdens on healthcare resources [45, 47]. We theorize that the prevalence of both of these STDs likely increase a county’s COVID-19 mortality rate directly and indirectly.

#### 4.1.6 Sleep, hospital beds/population, diabetes, cardiovascular disease, and exercise

Several other high-risk factors and determinants that were statistically insignificant by July 5 were observed to be significant by December 28. Percent of county with insufficient sleep (less than seven hours a night), hospital beds per population, prevalence of diabetes, and death rate cardiovascular disease were all associated with increased COVID-19 mortality (1.11, 1.10, 1.06, and 1.04 MRR, respectively). Meanwhile, percent of county with access to exercise opportunities was associated with a decrease in mortality (0.95 MRR). Diabetes and cardiovascular disease have been recognized as comorbidities, which may increase risk of severe complications from COVID-19 infection [26]. Regular exercise has been previously associated with positive health outcomes, and we observe that it continues to have a protective role in COVID-19 [17]. In contrast, insufficient sleep has been linked to an increased chance of severe chronic diseases like diabetes [58], and we find that this destructive effect may extend to COVID-19. The ratio of hospital beds per population is one measure of available healthcare resources, and a higher ratio is traditionally implicated with more urbanization [13, 61], so our results may suggest an increased risk of COVID-19 in urban environments, rather than a direct relationship between hospital beds availability and increased COVID-19 mortality rates.

#### 4.1.7 Other health-related factors

The other health-related factors we tested were statistically insignificant in the high-risk models and social determinant analyses in both time points. Infant mortality rate, ratio of primary care physicians to population, ratio of primary care clinicians other than physicians to population, prevalence of smoking, and prevalence of COPD appear to lack significance in aggregate data, despite some of these factors having literature about interactions with COVID-19 [26]. Including a large number of terms in the models may have diminished the effect of these factors. It is also possible that these factors may be regionally significant, and further analysis is needed at the sub-national level.

### 4.2 Socioeconomic Quality of Life

This section discusses the results presented in Figure 3.

#### 4.2.1 Disease Progression

The temporal variable of days since first infection has a high, statistically significant MRR of 2.23 for July 5 but was not observed to be significant by December 28. We likely observe that an early incidence of COVID-19 in a county is associated with the greatest increase in COVID-19 mortality, likely due to outlier metropolitan areas like New York City. It is also possible that as the pandemic progressed, the improved clinical experience of treating COVID-19, appropriate resources, and better compliance with social distancing is responsible for the statistically insignificant finding in December.

#### 4.2.2 Income

One standard deviation increase in county median income was associated with a 1.23 times increase in COVID-19 mortality (1.23 MRR, July 5). However, this changed to show a protective value of 0.95 MRR by December. In a traditional view of social determinants, higher income is associated with lower rates of disease [9]. One explanation for our initial finding between income and COVID-19 is that urban counties, where income is higher [8], were disproportionately affected by COVID-19 in the initial months. Over time, the traditionally implicated protective benefits likely had a greater impact on health outcomes, which is responsible for the protective MRR by December.

#### 4.2.3 Immigrant Communities

One standard deviation increase in percent of individuals not proficient in English was associated with a 1.49 increase in COVID-19 mortality in July. This decreased to a MRR of 1.07 by December. As of 2013, the percentage of Limited English Proficient (LEP) people who were foreign born was 81.3% [70]. Hence, these results suggest that immigrant populations are at higher risk from COVID-19, which is supported by previous work [22]. Historically, immigrants suffer from poverty, and many immigrants are barred from receiving healthcare programs such as Medicare in their first 5 years of living in the US. Additionally, undocumented immigrants are not eligible for any public programs. It is estimated that around 45% of documented immigrants and 65% of undocumented immigrants do not receive healthcare. This leaves the population much more vulnerable. LEP immigrants also receive lower quality of care and understanding of their medical condition due to language barriers in treatment [70]. Additionally, the observed decrease in MRR from 1.49 to 1.07 is consistent with the trend about the effects of socioeconomic disadvantage on COVID-19, as discussed above. Overall, immigrant populations are recognized to be at a higher risk for severe outcomes due to lower access and poorer quality of care.

#### 4.2.4 Poverty & Severe Housing Problems

One standard deviation increase in overcrowding, a measure of poor socioeconomic status [16], was associated with a 1.27 COVID-19 MRR by July 5. This variable is defined as the ratio of residences with more than one person per room to the total number of housing units in a county. Over-crowding is a metric of severe housing problems, previously associated with poor mental health outcomes [59], tuberculosis [59], and several other diseases [46]. Beyond signifying poor socioeconomic status, there is also physical risk with any infectious disease from overcrowding due to close proximity of potential carriers [5]. The MRR for overcrowding decreased to 1.06 by December 28, which is consistent with the decreasing trend in other explorations of socioeconomic disadvantage and COVID-19 in this study. However, despite the decreasing effect, the MRR of overcrowding in a county is still associated with a significant increase of COVID-19 mortality. In contrast, however, percent of county with severe housing burden, defined as families spending more than 50% of their income on housing, was negatively associated with COVID-19 mortality rate (0.95 MRR, December 28). Similarly, percent of county that is food insecure, defined as percent of population without adequate access to food, was also negatively associated with a COVID-19 mortality rate (0.93 MRR, December 28). Both of these social determinants are implicated in socioeconomically disadvantaged counties and are related to lower access to healthcare [50, 55]. One possibility is that systemic, socioeconomic intervention during the pandemic may be responsible for this finding, which some literature highlight as a possible method to combat the expected, negative effects of poverty [42].

#### 4.2.5 Air Pollution

One standard deviation increase in average daily air pollution was associated with a 1.37 increase in COVID-19 mortality rate in July 5, which decreased to 1.11 MRR by December 28. This finding supports Wu et al.’s results, indicating that counties with long-term air pollution are more likely to fall severely ill [68], although the impact of air pollution is observed to be decreasing over time and may need further study.

#### 4.2.6 Education

One standard deviation increase in percent of adults in county without high school education was associated with a 1.15 and 1.13 COVID-19 MRR in July and December, respectively. Average grade performance, defined as the outcomes of 3rd grade reading scores compared to the national average, was associated with a 0.86 MRR in July, but had no statistical significance by December. High school graduation rate in a county was associated with no statistical significance at either time point. Education is a well-studied determinant of adolescent health [64], and lower high school graduation has been specifically linked to poorer health outcomes in the past [39]. Moreover, grade performance is a metric for available educational opportunities [21] and a predictor of future health outcomes [31]. Our findings suggest that a county’s educational resources and quality, as reported by average grade performance and graduation rate, may have some protective effects in July, but have no measurable effects by December. However, the consistent, destructive relationship between the percent of adults without a high school education and COVID-19 still suggests that poor education can increase health risks.

#### 4.2.7 Motor Vehicle Mortality Rate and Long Commute

One standard deviation increase in motor vehicle mortality rate was associated with a 1.13 COVID-19 MRR on July 5. Increased motor vehicle mortality rate has been associated with counties with high uninsurance and high concentrations of ethnic minorities [36]. As discussed above, those with poor health care and certain minorities have an increased risk to COVID-19. Additionally, increased motor vehicle mortality rate is also associated with populations under the age of 18 [36], which we did find to be significantly associated with increased COVID-19. While this effect of motor vehicle mortality decreased to 1.05 MRR in December 28, we observed a statistically significant 0.96 MRR in December for percentage of county that drives alone and has a commute over 30 minutes. While this social determinant is traditionally associated increased obesity, less physical excercise, and worse mental health [52], we hypothesize that the transition to working from home may have a role in the protective MRR. While physical health benefits of working from home still need to be further elucidated, other benefits like mental health have been well-documented [44]. Overall, it is likely that the relationship between motor vehicle mortality rate, long commutes while driving alone, and COVID-19 has factors beyond the scope of this study, so future work is needed to fully understand this relationship.

#### 4.2.8 Social Association and Death Rate from All Causes

The relationships from county’s social association rate and cumulative death rate from all causes were both statistically insignificant in July but had significant COVID-19 MRRs of 1.09 and 1.11 by December, respectively. Although social association of a county through membership associations has been previously associated with positive health outcomes, increased social interaction may lead to a higher number of COVID-19 deaths [56]. Meanwhile, counties with high rates of all causes of mortality before the pandemic likely suffer from a mix of negative underlying living, healthcare, and working conditions. It is likely that the effects of both of these social determinants were statistically obscured by other factors in July, but the larger amount and more reliable data over time reveal their significance in December.

#### 4.2.9 Other Socioeconomic Factors

The other factors related to socioeconomic status that we tested were statistically insignificant in the high-risk models and social determinant associations at both times. These include percent of county that drives alone to work (without a long commute), percent of county that are homeowners, a county’s segregation index, death rate from assault, presence of water violation, unemployment rates, and days since mask required. As discussed above, including a large number of terms in the model may have diminished the effect of these factors and they may need further analysis at regional levels.

### 4.3 Race and Age Demographics

This section discusses the results presented in Figure 4.

#### 4.3.1 Race

Our results suggest that race was a better predictor of COVID-19 in July than in December. Counties experienced 1.25 and 1.18 MRR increases in COVID-19 mortality per standard deviation increase in Hispanic and Native American populations by July 5, respectively. These MRRs decreased to 1.11 and 1.07 by December 28, respectively. Additionally, while percent of African Americans and Asians in a county was associated with 1.39 MRR and 1.02 in July, there was no statistically significant relationship in December. Meanwhile, percent of county that is White was not statistically significant in either time point. Historically, ethnic minorities have disadvantaged access to healthcare, occupational opportunities, higher chronic stress, lower access to quality education, and more housing problems, which can contribute to health outcomes [27]. We conclude these racial inequalities have a role in the increased COVID-19 mortality rate, but they are observed to have a decreasing impact throughout the progression of the pandemic.

#### 4.3.2 Age

In contrast, we observe an increasingly strong relationship between age and COVID-19. While neither percent of county above 65 years old nor percent below 18 years old was significant by July, both terms were significant by December 28 (1.07 and 1.11 MRR, respectively). While other literature has recognized elderly populations as at risk [26], we additionally find that an increase in youth in a county is associated with an increase in COVID-19 mortality. Youth have been suspected to be a source of transmission [20], which may be responsible for this finding.

## 5 CONCLUSION

Overall, we identify that counties with Hispanic and Native American ethnic minorities, populations under 18 and above 65 years of age, worse healthcare resources, higher social association, higher prevalence of comorbidities like pediatric asthma and diabetes and cardiovascular disease, more overcrowding, higher motor vehicle mortality, earlier exposure to COVID-19, higher vaccination rates and mammogram screenings among Medicare FFS users, and higher air pollution continue to have an increased risk of COVID-19 mortality. Like other diseases [14], we recognize that the severity of COVID-19 is greatly worsened by socioeconomic inequality. Additionally, the temporal trends observed in this study suggest that the policies implemented during this pandemic may have been effective, as the impact of several socioeconomic metrics (overcrowded housing, immigrant populations, poor health in a county) appear to significantly decrease over time. However, future analysis is needed to further support this finding, and to additionally explore the decreasing impacts of air pollution and income, and the increasing impact of age demographics.

Care must be taken in contextualizing these results due to inherent limitations of county-level ecological studies. The relationships observed in aggregate data cannot be translated from the county level to individuals from an ecological study alone. Rather, these studies identify useful trends and hypotheses for future analysis at the individual level [63]. Accounting for possible confounds is also essential in ecological studies, which we achieve by including several controls in every model. Other limitations of the study stem from the availability and accuracy of the data. Literature suggests that both the reported counts of infection and the reported COVID-19-related deaths have inconsistencies [12, 57], and the lack of proper testing has also been highlighted [33]. Moreover, changes in reporting measures and standards of care over the study period may result in a regression to the mean, which should also be considered in temporal analysis.

As more detailed information becomes available, it would prove useful for future study to verify our results using clinical and individual-level data. Further investigation of the relationship between COVID-19 and adult chronic lung disease may be especially valuable, as there is currently varying evidence. Additionally, the relationships between COVID-19, poverty, air quality, income, age, motor vehicle mortality rate, vaccination rate, breast cancer screening rate, mental health, and alcohol consumption need to be elucidated in more detail. These analyses may reveal further interactions between social determinants and COVID-19 mortality.

## Data Availability

A publicly-accessible repository containing all data used in this work has been available and is cited in the paper. All data used in this work was obtained via public sources.

https://github.com/TheRensselaerIDEA/COVIDMINDER/tree/master/social-determinants-paper

## ACKNOWLEDGMENTS

This study was supported by the Rensselaer Institute for Data Exploration and Applications, the Data INCITE Lab, and United Health Foundation grant 1990. Additionally, thank you to Ariella D. Sprague, Hongxi Mou, Tiburon L. Benavides, Sarah M. Ahn, and Cole A. Reschke for their contributions to an earlier version of this work.

## Footnotes

1 https://github.com/TheRensselaerIDEA/COVIDMINDER/tree/master/social_determinants_paper

